# Impact of loss-of-function in angiopoietin-like 4 on the human phenome

**DOI:** 10.1101/2024.02.13.24302776

**Authors:** Eloi Gagnon, Jérome Bourgault, Émilie Gobeil, Sébastien Thériault, Benoit J. Arsenault

**Affiliations:** Centre de recherche de l’Institut universitaire de cardiologie et de pneumologie de Québec, Québec (QC), Canada; Department of Medicine, Faculty of Medicine, Université Laval, Québec (QC), Canada; Department of Molecular Biology, Medical Biochemistry and Pathology, Faculty of Medicine, Université Laval, Québec (QC), Canada

## Abstract

**Background:** Carriers of the E40K loss-of-function variant in Angiopoietin-like 4 (*ANGPTL4*), have lower plasma triglyceride levels as well as lower rates of coronary artery disease (CAD) and type 2 diabetes (T2D). These genetic data suggest ANGPTL4 inhibition as a potential therapeutic target for cardiometabolic diseases. However, it is unknown whether the association between E40K and human diseases is due to linkage disequilibrium confounding. The broader impact of genetic ANGPTL4 inhibition is also unknown, raising uncertainties about the safety and validity of this target.

**Methods:** To assess the impact of ANGPLT4 inhibition, we evaluated the impact of E40K and other loss-of-function variants in *ANGPTL4* on a wide range of health markers and diseases.. The E40K impact was assessed in 29 publicly available genome-wide association meta-analyses of European ancestry on cardiometabolic traits and diseases as well as 1589 diseases assessed in electronic health record within FinnGen (n=309,154). To ensure that these relationships were truly causal, and not driven by other correlated variants, we used the Bayesian fine mapping algorithm CoPheScan.

**Results:** The CoPheScan posterior probability of E40K being the causal variant for triglyceride was 99.99%, validating the E40K to proxy lifelong lower activity of ANGPTL4. The E40K mutation was associated with lower risk of CAD (odds ratio [OR]=0.84, 95% CI=0.81 to 0.87, p=3.6e-21) and T2D (OR=0.91, 95% CI=0.87 to 0.95, p=2.8e-05) in GWAS meta-analyses, with results replicated in FinnGen. These significant results were also replicated using other rarer loss-of-function variants identified through whole exome sequencing in 488,278 participants of various ancestry in the UK Biobank. Using a Mendelian randomization study design, the E40K variant effect on cardiometabolic diseases was concordant with lipoprotein lipase enhancement (r=0.85), but not hepatic lipaseenhancement (r=-0.10), suggesting that ANGPTL4 effects on cardiometabolic diseases are potentially mainly mediated through lipoprotein lipase. The E40K variant did not significantly increase the risk of any of the 1589 diseases tested in FinnGen.

**Conclusion:** ANGPTL4 inhibition may represent a potentially safe and effective target for cardiometabolic diseases prevention or treatment.

## Introduction

Angiopoietin-like 4 (ANGPTL4) plays a key role in lipoprotein-lipid metabolism and energy patitioning. ANGPTL4 inhibits lipoprotein lipase (LPL) mediated lipoprotein uptake by adipose tissue leading to a decrease in circulating lipoproteins and lipids (Kersten 2021). Carriers of the E40K loss-of-function variant in *ANGPTL4* are characterized by lower plasma triglyceride levels as well as lower rates of coronary heart disease (CAD) and type 2 diabetes (T2D) (Dewey et al. 2016; Gusarova et al. 2018). These genetic data suggest that ANGPTL4 inhibition may become a pharmacological strategy of interest in the prevention or treatment of cardiometabolic diseases, especially in individuals with elevated triglycerides levels. For ANGPTL4 to be a viable target, its inhibition must be proven safe. Concerns about the safety of ANGPTL4 inhibition arise from the observation that monoclonal antibody-induced ANGPTL4 inhibition in mice and monkeys led to the accumulation of lipid in mesenteric lymph nodes (Dewey et al. 2016). Consequently, the safety of ANGPTL4 inhibition in humans remains uncertain.

A powerful method to understand the functional impact of proteins such as ANGPTL4 is to leverage their loss-of-function variants. Genetic variations are randomly distributed during gametogenesis and remain constant throughout life, making their associations with diseases unlikely to be influenced by environmental confounding factors or reverse causality, hence likely to be causal. The most common functional variant in ANGPTL4 is E40K, which substitutes a glutamic acid for a lysine giving rise to an unstable ANGPTL4 protein (Yin et al. 2009). The frequency of E40K carriers varies from 0.1% in individuals of African ancestry, to approximately 3% in individuals of European ancestry to 18% in a Tunisian population (Abid et al. 2016). The well-replicated associations of E40K with lipid levels and diseases could be spurious (i.e., non-causal) if driven by another correlated variant. In fact, the previous cohort studies on E40K carriers were unable to distinguish true causal effects from confounding due to linkage disequilibrium (Myocardial Infarction Genetics and CARDIoGRAM Exome Consortia Investigators et al. 2016; Dewey et al. 2016; Gusarova et al. 2018). To address linkage disequilibrium confounding, it is essential to employ fine mapping methods such as CoPheScan (Coloc adapted Phenome-wide Scan) (Manipur et al. 2023). CoPheScan is a Bayesian approach that integrates the linkage disequilibrium matrix and information on the neighbouring variants to address linkage disequilibrium confounding (Manipur et al. 2023).

To gain insight into the effects of ANGPTL4 inhibition, we evaluated the effect of the E40K variant on 29 cardiometabolic traits and 1589 human diseases, while addressing confounding due to linkage disequilibrium. We show that loss-of-function in *ANGPTL4*, proxying lifelong inhibition of ANGPTL4, lowers the risk of CAD, T2D and aortic stenosis, and does not significantly increase the risk of any of the 1589 diseases tested.

## Methods

### Genome-wide association studies of cardiometabolic traits and human diseases

The GWAS for cardiometabolic traits and diseases, all publicly available, were selected based on the sample size and cases/controls ratio to maximize statistical power (Supplementary Table 1). Specifically there were 20 cardiometabolic traits and 9 cardiometabolic diseases (Richardson et al. 2020; Sinnott-Armstrong et al. 2021; Graham et al. 2021; Aragam et al. 2022; Mishra et al. 2022; Mahajan et al. 2022; Liu et al. 2021; Ward et al. 2021). For non-alcoholic fatty liver disease (NAFLD), we meta-analyzed GWAS summary statistics from Estonian Biobank, deCODE genetics, UKB, INSPIRE+HerediGene (USA), FinnGen and eMERGE networks (16,532 cases and 1,240,188 controls) (Ghodsian et al. 2021; Sveinbjornsson et al. 2022). For estimated glomerular filtration rate (eGFR), we used a GWAS meta-analysis of CKDGen and UK Biobank (n=1,004,040) (Stanzick et al. 2021). These summary statistics were reported in log(eGFR). For better interpretability and comparability, we transformed them to a one standard deviation (SD) scale using the sdY.est function in the coloc package (Wallace 2021). The other continuous variables were already inverse-rank normal transformed in the original GWAS.

We included all diseases with over 1000 cases in FinnGen data freeze 7 totalling 1589 distinct diseases established with electronic health record ICD10 codes (Supplementary Table 1). FinnGen is a population-based cohort totalling 309,154 genotyped individuals of European ancestry (Kurki et al. 2023). Given the relatively high median age of participants (63 years) and the substantial fraction of hospital-based recruitment, FinnGen is enriched for disease end-points. All end-points were adjusted for sex, age, genotyping batch and ten first principal genetic components as covariates prior to GWAS. All GWAS were performed using the SAIGE (v.0.35.8.8) algorithm (Zhou et al. 2018).

### Genetic colocalisation

To test for confounding due to linkage disequilibrium, we used the CoPheScan algorithm. CoPheScan is a Bayesian approach that integrates the linkage disequilibrium matrix and information on the neighbouring variants to address linkage disequilibrium confounding.. We used CoPheScan to help distinguish the E40K associations with traits that might be confounded by linkage disequilibrium from those that are true effects. We included all SNPs within 300 Kb upstream or downstream of the *ANGPTL4* gene region and systematically tested whether the E40K was the likely causal variant for all tested traits. CoPheScan priors were computed using a hierarchical Bayesian model. Bayes factors were calculated using the SuSIE algorithm allowing for multiple causal variants. SuSIE was computed with a LD matrix obtained using the 1000-genome reference panel of individuals of European ancestry (Zou et al. 2022). CoPheScan returns posterior probability for three mutually exclusive hypotheses: H_n_=No association with the query trait; H_a_=Causal association of a variant other than the query variant with the query trait; and H_c_=Causal association of the query variant with the query trait. A high H_c_ supports that the association is not due to linkage disequilibrium.

We investigated shared genetic etiology across lipids and cardiometabolic diseases using multi-trait colocalization analyses with the HyPrColoc R package (Foley et al., 2021). HyPrColoc extends traditional genetic colocalization methods by estimating posterior probability of more than two traits sharing the same causal variants. If the traits do not share a causal SNP, a branch-and-bound selection algorithm is employed to highlight a subset of traits that share a causal SNP. We used the default uniform prior for the analysis. Prior.1 - the prior probability of a SNP being associated with one trait-was set to 1e-4 and prior.c - the probability of a SNP being associated with an additional trait given that the SNP is associated with at least one other trait-was set to 0.02. HyPrColocalization analyses are sensitive to the choice of priors. The priors were therefore iteratively incremented to assess the stability of clusters as a sensitivity analysis. All analyses were performed with the genetic region spanning 300 Kb around the *ANGPTL4* gene region. The colocalization events were visualized with regional stack plot created with the *gassocplot* package.

### ANGPTL4 loss-of-function mutations in UK Biobank

Access to the UK Biobank was granted under UK Biobank application 25205. The UK Biobank is a population-based cohort of approximately 500,000 participants, aged 40 to 69 years recruited during 2006–2010 from several centres across the UK. We studied a subsample of 488,278 individuals from mixed ancestries with whole exome sequence available. These sequences were annotated with the Ensembl Variant Effect Predictor (McLaren et al., 2016) to identify loss-of-function variants in the *ANGPTL4* gene region. Loss-of-function variants were defined as either frameshift, missense, splice, nonsense, start-lost or stop-lost variants and grouped according to the Phred-scaled Combined Annotation Dependent Depletion (CADD) score (Rentzsch et al., 2021). Noncarriers of these variants were assigned to the control group, carriers with at least one variants with CADD score > 20 and significantly associated with triglycerides (p<0.05) were assigned to the carrier group, individuals with loss-of-function mutation CADD < 20 or not associated with triglycerides were excluded. The E40K variant CADD score was 31 and it was associated with triglyceride levels at p<7e-136. To specifically assess the impact of other loss-of-function mutations in *ANGPTL4*, the analyses were performed by including and excluding E40K carriers.

We defined CAD, T2D and aortic stenosis using Hospital Episode Statistics ICD10 and OPCS4 codes. CAD cases were defined as participants with ICD-10 codes for MI (I21.X, I22.X, I23.X, I24.1, or I25.2), other acute ischemic heart diseases (I24.0, I24.8-9) or chronic ischemic heart disease (I25.0-25.1, I25.5-25.9), OPSC-4 codes for coronary artery bypass grafting (K40.1-40.4, K41.1-41.4, K45.1-45.5), for coronary angioplasty, with or without stenting (K49.1-49.2, K49.8-49.9, K50.2, K75.1-75.4, K75.8-75.9). AS cases were defined as participants with aortic valve stenosis (I35.X). Participants with rheumatic fever (I01-9) prior to AS diagnosis were excluded from the analysis on AS. T2D cases were defined as participants with type 2 diabetes mellitus (E11, E12, E13, E14).

To assess the association of loss-of-function mutation on the incidence of diseases, we performed Cox regression. All models were adjusted for variables at recruitment including age at recruitment, sex, and the first 10 principal components of ancestry. To model the lifelong effects of carrier status, the follow-up was started at birth and ended at censoring, death or the date of the data release (2021-11-12). Results are presented as estimated hazard ratios (HR) for incident diagnosis, with 95% confidence intervals.

### Effect of LPL and hepatic lipase (HL) on cardiometabolic traits and diseases

To gain insights into the mechanism linking genetic ANGPTL4 inhibition to diseases, we investigated the effects of LPL and HL, two enzymes inhibited by ANGPTL4, on cardiometabolic traits and diseases. LPL enhancement was proxied using all independent variants (r^2^<0.1) 300 Kb downstream and upstream of the *LPL* gene region significantly (pval <5e-8) associated with triglyceride levels (Graham et al. 2021). Similarly, HL enhancement was proxied using all independent variants (r^2^<0.1) 300 Kb downstream and upstream of the *LIPC* gene region significantly (pval<5e-8) associated with triglyceride levels. Harmonization was performed by aligning the effect sizes of different studies on the same effect allele. When a particular SNP was not present in the outcome datasets, we used a proxy SNPs (*r^2^*> 0.8) obtained using linkage disequilibrium information from the 1000 Genomes Project European samples.

Instrument strength was assessed using the F-statistic (Burgess, Thompson, and CRP CHD Genetics Collaboration 2011), and the variance explained assessed using the *r^2^* (Pierce, Ahsan, and VanderWeele 2011). The effect of each gene on cardiometabolic traits and diseases was calculated using the inverse variance weighted method with multiplicative random effects (Burgess, Foley, and Zuber 2018) when the number of genetic instruments were over one, or the Wald ratio in the case of the E40K because the number of instruments was equal to one. We compared the effect of LPL and HL on cardiometabolic traits and diseases to the ANGPTL4 inhibition on these same cardiometabolic traits and diseasesu using a Pearson’s correlation coefficient.

## Results

The E40K variant was associated with a more favorable lipoprotein-lipid profile, including lower triglyceride and apolipoprotrein (apo) B levels, higher HDL cholesterol levels, but was not associated with LDL cholesterol levels (Figure 1). Additionally, it was associated with lower waist-to-hip ratio, but not with BMI, indicating a more favourable body fat distribution in carriers. Information on the GWAS summary statistics used are presented in Supplementary Table 1. Overall, the E40K showed favorable or neutral associations with the 20 cardiometabolic traits tested (Supplementary Table 2).

**Figure 1.**
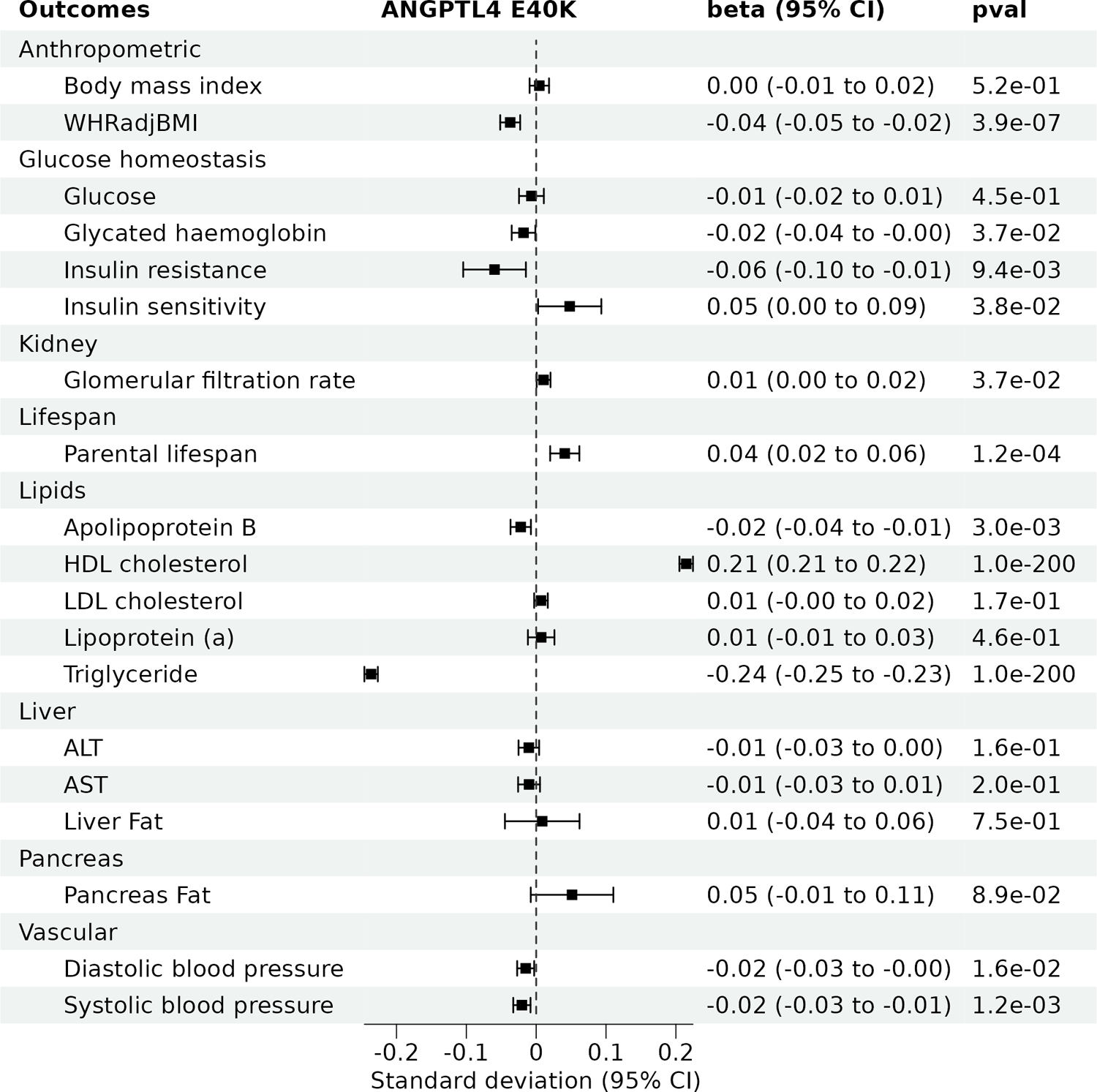
Impact of the *ANGPTL4* loss-of function variant E40K on cardiometabolic traits. All cardiometabolic traits have a SD of one. ALT = alanine aminotransferase, AST = aspartate aminotransferase, BMI = body mass index, HDL = high-density lipoprotein, LDL = low-density lipoprotein, WHRadjBMI = waist-to-hip ratio adjusted for body mass index. CI = confidence interval

The E40K variant was protective for several cardiometabolic diseases including aortic stenosis (odds ratio [OR] = 0.82, 95% CI=0.74 to 0.92, p=7.1e-04), CAD (OR = 0.84, 95% CI=0.81 to 0.87, p=3.6e-21) and T2D (OR = 0.91, 95% CI=0.87 to 0.95, p=2.8e-05) (Figure 2). The E40K was also associated with lower risk of stroke, ischemic stroke and heart failure. There was no evidence for an association with acute pancreatitis, chronic kidney disease, or non-alcoholic fatty liver disease. The E40K was also associated with longer parental lifespan, indicating a potentially favourable benefit/risk balance (Figure 1). In summary, the E40K variant was either protective or neutral of all cardiometabolic diseases tested.

**Figure 2.**
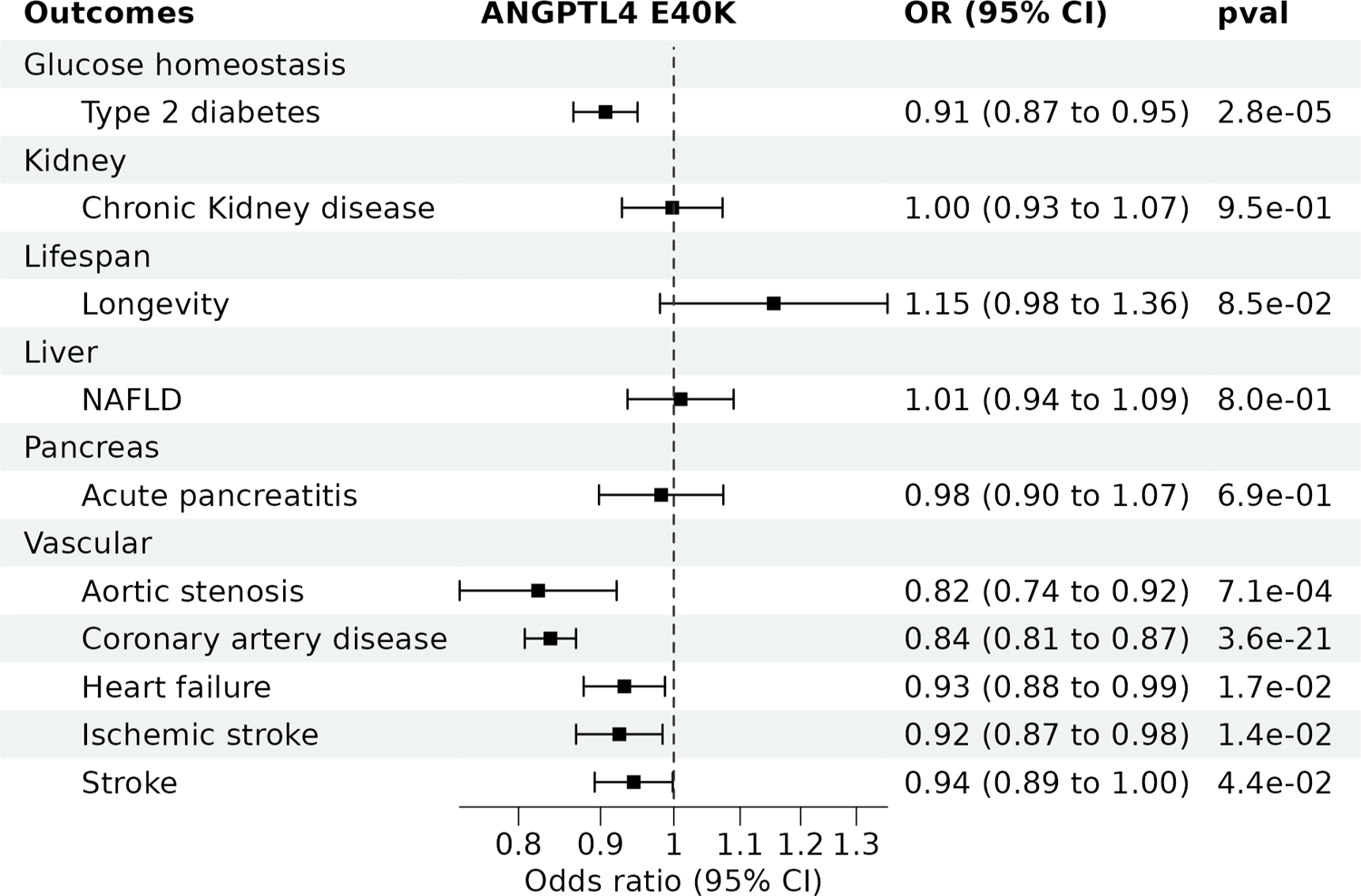
Impact of the *ANGPTL4* loss-of function variant E40K on cardiometabolic diseases. NAFLD = non-alcoholic fatty liver disease. CI = confidence interval and OR = Odds ratio.

Other less frequent loss-of-function mutations in *ANGPTL4* were also associated with healthier lipid levels and decreased risk of diseases (Figure 3). By scanning whole-exome sequence of UK biobank participants, we identified 2835 carriers (excluding E40K) or 20,015 carriers (including E40K) of deleterious *ANGPTL4* variants (CADD score > 20 and associated with triglycerides with a p-value<0.05). *ANGPTL4* loss-of-function carriers had lower levels of triglycerides, higher levels of HDL cholesterol and lower incidence of T2D (Figure 3). When including E40K carriers, loss-of-function in ANGPTL4 was associated with lower CAD incidence (Figure 3 panel B). When excluding E40K carriers, loss-of-function in *ANGPTL4* was associated with lower CAD incidence but the association did not reach statistical significance (HR = 0.90 95% CI=0.80 to 1.02, p=9.9e-02). Loss-of-function in *ANGPTL4* was associated with lower aortic stenosis incidence only when including E40K carriers. The directionally consistent associations of E40K and other rarer loss-of-function variant in *ANGPTL4* support the role of ANGPTL4 in CAD and T2D.

**Figure 3.**
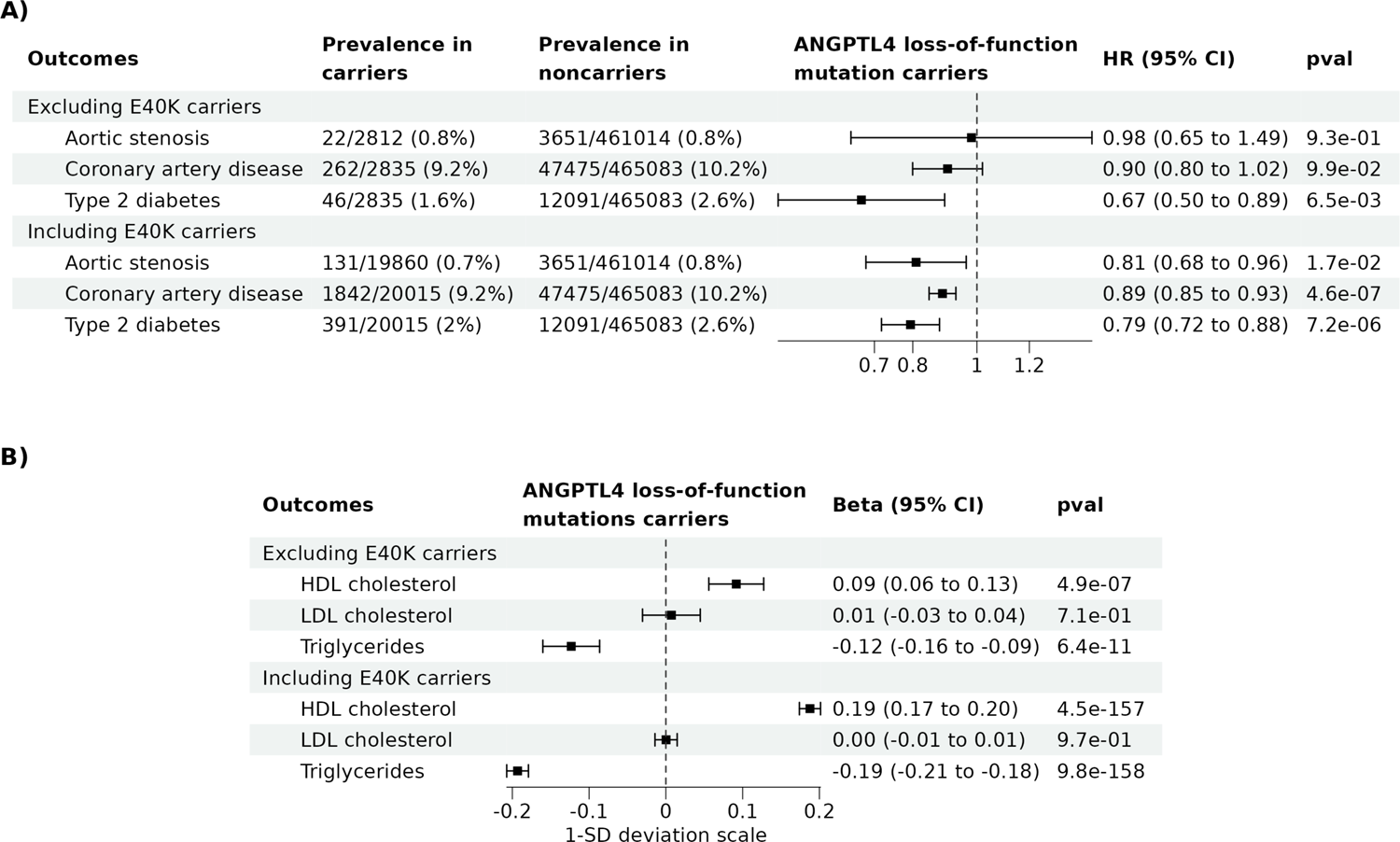
Health impact of *ANGPTL4* loss-of function mutations in the UK Biobank. Impact of *ANGPTL4* variants (including E40K and excluding E40K) on A) incidence of coronary artery disease, type 2 diabetes and aortic stenosis and B) high-density lipoprotein (HDL) cholesterol, low-density lipoprotein (LDL) cholesterol and triglyceride levels.

The E40K protective effects on diseases were not due to confounding by linkage disequilibrium as demonstrated by colocalization algorithms. Within the *ANGPTL4* gene region +/- 300 Kb, CoPheScan and HyPrColoc prioritised E40K as the unique causal variant. For example, although the CoPhesScan posterior probability of E40K being the unique causal variant was 0.06% for AS, this probability was 98% for T2D and 99.99 % for triglyceride levels, HDL cholesterol levels, and CAD (Supplementary Table 3). These high probabilities support that E40K associations are causal and unaffected by linkage disequilibrium confounding. Similarly, the HyPrColoc algorithm showed a posterior probability of multi-trait colocalization of 0.79 for HDL cholesterol levels, triglyceride levels, T2D, CAD and AS (Figure 4). Notably, the rs116843064-A (i.e., the E40K) explained 100% of the posterior probability of multi-trait colocalization, providing evidence for E40K as a shared causal variant. In summary, both colocalization approaches consistently validated E40K as a reliable genetic proxy for lifelong ANGPTL4 inhibition.

**Figure 4.**
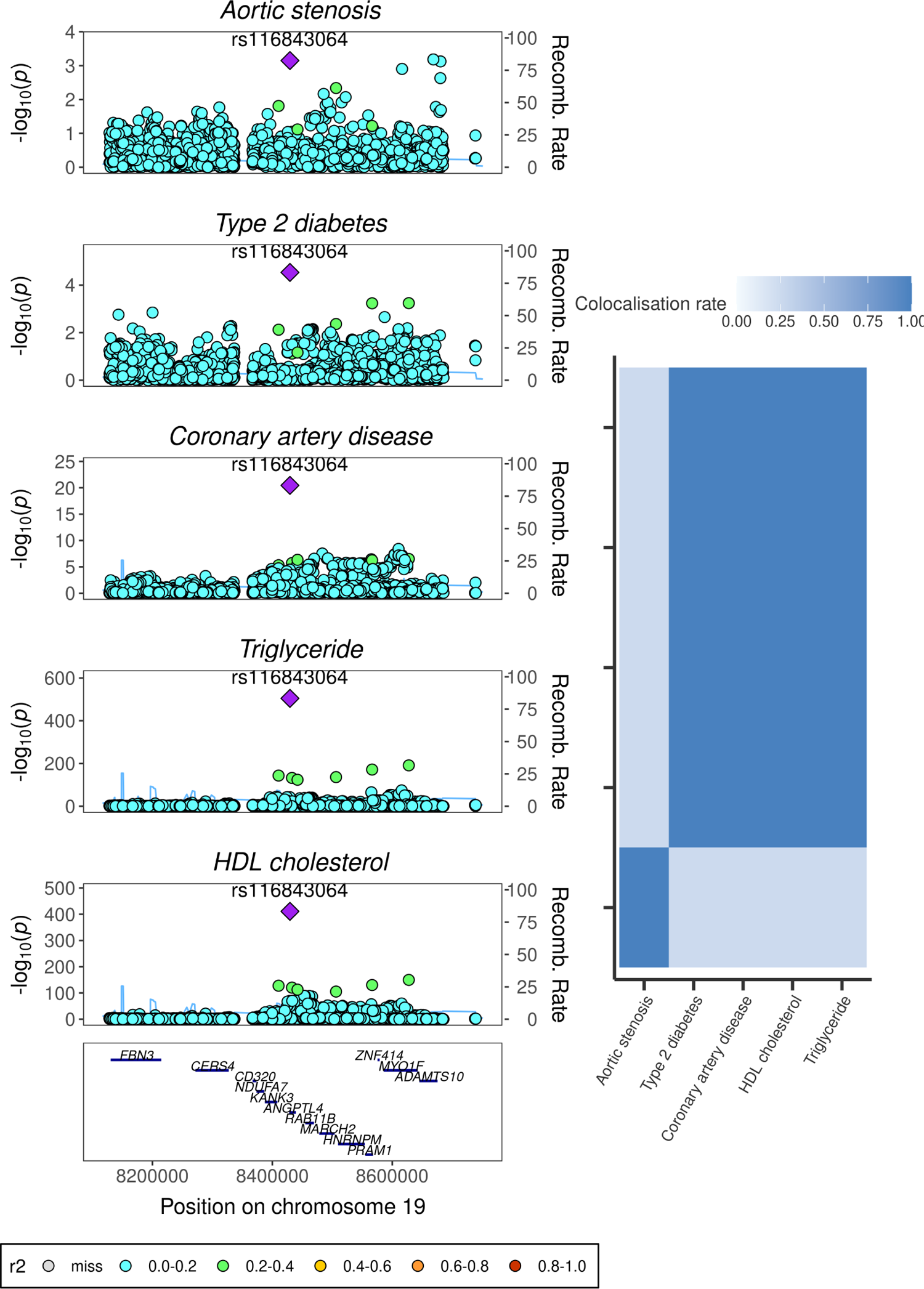
Multi-trait genetic colocalization for lipids and cardiometabolic diseases. The left panel shows the regional association plot with the highlighted genetic variant prioritized by HyPrColoc. The right panel shows the heatmap of the proportion of colocalization on different range of prior values.

### Phenome-wide study of ANGPTL4 E40K in FinnGen

To assess the safety of ANGPTL4 inhibition, we examined the impact of E40K on 1589 diseases in FinnGen. E40K was associated with a lower risk of aortic stenosis, CAD and T2D, replicating our initial results obtained through GWAS meta-analysis (Figure 5). The E40K variant was not associated with lymphadenopathy, ascites, or peritonitis, as observed in whole-body *ANGPTL4* knockout mice. Notably, the E40K did not significantly increase the risk of any of the other diseases tested suggesting that the benefits of genetic inhibition of *ANGPTL4* may have more health advantages than consequences (Supplementary Table 2).

**Figure 5.**
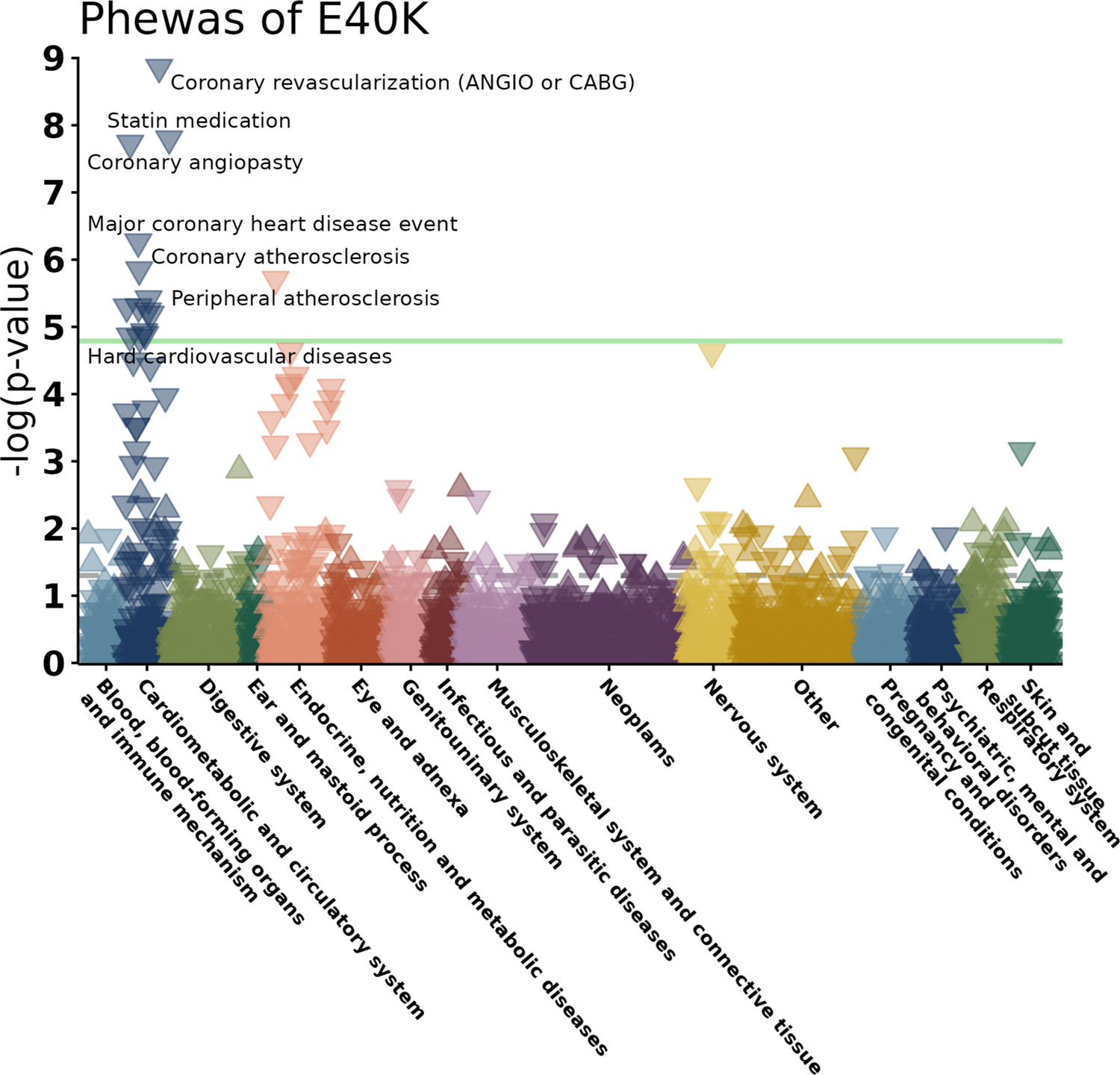
Phenome-wide association study of the *ANGPTL4* loss-of function variant E40K on 1589 diseases in FinnGen. The green line is the threshold for multiple testing significance. Triangles pointing down represent negative associations (i.e., E40K lowers the risk of the disease), whereas triangles pointing up represent positive associations (i.e., E40K increases the risk of the disease). Associations that pass multiple testing correction are annotated and encompass atherosclerotic conditions and type 2 diabetes. Among the triangles pointing up, all are below the green significance line, meaning that E40K is not significantly associated with an increase risk of diseases.

### Exploring E40K mechanisms

The impact of ANGPTL4 on diseases is likely tied to its inhibition of enzymes such as LPL andHL. To shed light on this mechanism, we assessed the effect of LPL and HL on cardiometabolic traits and diseases and compared their effects with those resulting from ANGPTL4 inhibition (Supplementary Table 4). LPL and HL enhancement were proxied with independent SNPs in their respective gene region +/- 300 Kb significantly associated (pval<5e-8) with triglycerides. Specifically LPL enhancement was proxied with 71 independent SNPs in the LPL region +/- 300 Kb significantly associated with triglycerides (r^2^ = 1.92%; F-statistics = 343) and HL enhancement was proxied with 23 SNPs in the region of the gene encoding HL significantly associated with triglyceride levels (r^2^ = 0.24%; F-statistics = 131). Genetically predicted enhancement of LPL and ANGPTL4 inhibition had similar effects on cardiometabolic traits and diseases (Pearson’s correlation = 0.85) (Figure 6). By contrast, ANGPTL4 and HL had distinct associations with cardiometabolic traits and diseases (Pearson’s correlation = −0.10). These data suggest that ANGPTL4 effects on cardiometabolic traits and diseases are potentially mainly mediated through its effect on LPL, but not through its effect on HL.

**Figure 6.**
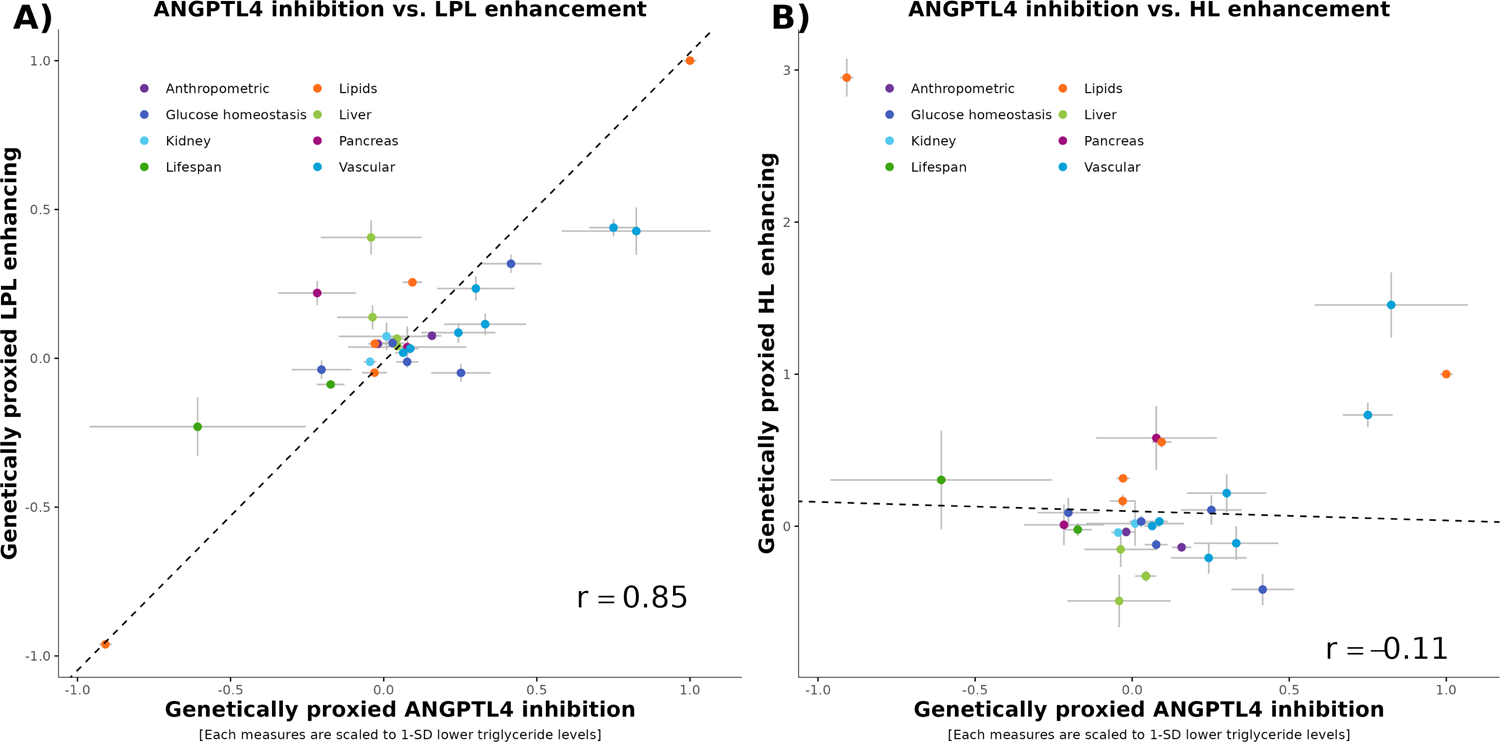
Impact on genetic inhibition of *ANGPTL4* on cardiometabolic traits and diseases in relation with the effect on genetic enhancement of lipoprotein lipase and hepatic lipase on the same traits and diseases. ANGPTL4 inhibition was proxied with the E40K. Lipoprotein lipase (LPL) and hepatic lipase (HL) enhancing were proxied with variants from their respective gene region significantly associated with triglyceride levels. Each effect is scaled to a one standard deviation effect on triglyceride levels. Panel A) ANGPTL4 inhibition vs. LPL enhancement. Panel B) ANGPTL4 inhibition vs. HL enhancement.

## Discussion

To explore the effects of genetic inhibition of *ANGPTL4* on human health, we performed a systematic assessment of loss-of-function in *ANGPTL4* and their associations with cardiometabolic traits and human diseases. The E40K, the most common loss-of function variant in *ANGPTL4*, was protective for CAD and T2D, as suggested by previous reports, as well as protective for aortic stenosis,. These associations were not confounded by linkage disequilibrium, consistent with a true causal effect of *ANGPTL4* genetic inhibition. Importantly, the E40K was not significantly associated with an increase in any of the 1589 diseases tested.

Altogether, these results provide robust genetic evidence that ANGPTL4 may represent a safe and effective therapeutic target to prevent or treat cardiometabolic diseases. Our phenome-wide association study showed that the E40K variant lowered risk of T2D, CAD and AS, but did not increase the risk of any of the 1589 tested, supporting ANGPTL4 inhibition as a viable therapy. Both E40K and other rarer loss-of-function mutations in *ANGPTL4* were associated with lower triglyceride levels and higher HDL cholesterol levels. Similar perturbations of lipid levels have been observed in mice with *ANGPTL4* whole-body knockout (Lichtenstein et al. 2010), liver-specific knockout (Singh et al. 2021) and adipose tissue-specific knockout (Aryal et al. 2018), providing external validation for our use of the E40K to proxy ANGPTL4 inhibition. The inhibition of ANGPTL4 seemed to be associated with lymphadenopathy, ascites, and peritonitis in mice (Lichtenstein et al. 2010) and monkeys (Dewey et al. 2016). By contrast, the ANGPTL4 E40K variant did not significantly increase the risk of these diseases, supporting the safety of ANGPTL4 inhibition in humans. The safety of human ANGPTL4 inhibition is further supported by a recent randomized, double-blind, placebo-controlled Phase I study evaluating the safety of a GalNAc conjugated ASO against ANGPTL4. The treatment did not cause serious adverse events but caused mild resolving injection site reactions (89% of treated, 0% of placebo) (K. Nilsson 2023).

Our results show that genetically predicted inhibition of ANGPTL4 and genetically predicted enhancement of LPL have very similar effects on cardiometabolic diseases (*r*=0.85), suggesting that the effects of ANGPTL4 on cardiometabolic diseases are potentially mainly mediated through LPL. In support of this notion, ANGPTL4 was shown to bind and inhibit LPL (Gutgsell et al. 2019). Furthermore, the E40K proxying inhibition of ANGPTL4 and rs115849089_A proxying enhancement of the activity of LPL were shown to have very similar effects on metabolites (r2>0.96), strongly suggesting that ANGPTL4 impacts plasma lipids exclusively via LPL (Wang, n.d.; Landfors, Chorell, and Kersten 2023). ANGPTL4 could also antagonise HL, as mice with hepatocyte-specific knock of ANGPTL4 have higher HL activity (Singh et al. 2021). Despite the potential inhibitory effect of ANGPTl4 on HL, we did not find evidence that ANGPTL4 influenced cardiometabolic disease through HL. In summary, our results and those of others (Landfors, Chorell, and Kersten 2023) support that ANGPTL4 effects on diseases are related to the activity of *LPL* and not *LIPC*, the gene encoding HL.

Our genetic data shows that loss-of-function variants in ANGPTL4 have no effect on lipoprotein(a), no effect on LDL cholesterol, a weak effect on ApoB and a stronger effect on CAD. These genetic data suggest that ANGPTL4 inhibition protects from CAD in part from ApoB lowering, but also through other mechanisms. The major effect of ANGPTL4 inhibition is triglyceride lowering, which has been initially argued as an independent causal factor for CAD (Gotto 1998). However, this notion has recently been challenged by high throughput multivariable Mendelian randomization studies showing no effect of triglyceride when ApoB is taken into account (Zuber et al. 2021). Genetic data support that the clinical benefit of lowering TG and LDL-C levels is proportional to the absolute change in ApoB (Ference et al. 2019), further emphasizing ApoB as the main risk factor. Hence, genetic inhibition of ANGPTL4 seems to reduce CAD risk not only through its lowering of ApoB-containing particle, but also possibly though other mechanism. For example, our data and those of others (Aryal et al. 2018) show that ANGPTL4 inhibition promotes fat storage at the periphery instead of at the abdomen, which protects from atherosclerotic diseases (Després 2012). ANGPLT4 inhibition could also improve insulin sensitivity, which could reduce coronary artery disease risk. Altogether, this data shows that ANGPTL4 inhibition may protect from CAD probably through multiple mechanisms, including ApoB lowering, insulin sensitivity and a more favourable body fat distribution.

The integration of these results supports that the inhibition of ANGPTL4 activates LPL which ultimately lowers CAD risk through mechanisms that are partly independent of ApoB lowering. Yet, the specific tissue of action remains uncertain as ANGPTL4 loss-of-function mutations inhibit ANGPTL4 in all tissues. ANGPTL4 is abundantly expressed in numerous human tissues, including kidney, intestine, and heart, but most abundantly in adipose tissue and next in liver (THE GTEX CONSORTIUM 2020). By contrast, LPL and HL are more tissue specific. LPL is expressed 1621 times more in the adipose tissue compared to the liver, whereas the LIPC is expressed 148 times more in the liver compared to the adipose tissue (THE GTEX CONSORTIUM 2020). We have shown that ANGPTL4 effects on diseases may be related to adipose tissue-enriched LPL and not to the liver-enriched LIPC. Therefore, it seems likely that ANGPTL4 acts on diseases primarily through its action in the adipose tissue. Ultimately, whether liver-specific inhibition of ANGPTL4 can increase LPL activity will be settled by future phase 2 trial studying liver targeted ASO on ANGPTL4 (Lipigon Pharmaceuticals 2023).

In conclusion, these results support that inhibition of ANGPTL4 may have considerable health benefits such as a reduction in the risk of CAD, T2D and aortic stenosis while possibly increasing lifespan. These effects may be mostly due to the activation of LPL. ANGPTL4 inhibition may represent a potentially safe and effective target for cardiometabolic diseases prevention or treatment.

## Data Availability

All data used in this study are in the public domain. Supplementary Table 1 describes the data used and relevant information to retrieve the summary statistics.

## Declarations

### Institutional Review Board Approval

All data used in this study are in the public domain. All participants provided informed consent and study protocols were approved by their respective local ethical committees. This project was approved by the Institutional Review Board of the Quebec Heart and Lung Institute.

### Code Availability

Code to reproduce the results of this manuscript is available on https://github.com/gagelo01/ANGPTL4

## Funding

EG holds a doctoral research award from the *Fonds de recherche du Québec: Santé*. (FRQS). JB holds a masters research award from the FRQS. ÉG holds a Doctoral Research Award from the Canadian Institutes of Health Research. BJA holds a senior scholar awards from the FRQS.

### Authors’ contributions

Data acquisition and analysis EG, JB, ÉG and ST. Conception and design EG, BJA. Drafting of the work EG, BJA. All authors approved the final version of the manuscript.

## References

Abid, Kaouthar, Thouraya Trimeche, Donia Mili, Mohamed Amine Msolli, Imen Trabelsi, Semir Nouira, and Abderraouf Kenani. 2016. ‘ANGPTL4 Variants E40K and T266M Are Associated with Lower Fasting Triglyceride Levels and Predicts Cardiovascular Disease Risk in Type 2 Diabetic Tunisian Population’. Lipids in Health and Disease 15 (March): 63. 10.1186/s12944-016-0231-6.

Aragam, Krishna G., Tao Jiang, Anuj Goel, Stavroula Kanoni, Brooke N. Wolford, Deepak S. Atri, Elle M. Weeks, et al. 2022. ‘Discovery and Systematic Characterization of Risk Variants and Genes for Coronary Artery Disease in over a Million Participants’. Nature Genetics 54 (12): 1803–15. 10.1038/s41588-022-01233-6.

Aryal, Binod, Abhishek K. Singh, Xinbo Zhang, Luis Varela, Noemi Rotllan, Leigh Goedeke, Balkrishna Chaube, et al. 2018. ‘Absence of ANGPTL4 in Adipose Tissue Improves Glucose Tolerance and Attenuates Atherogenesis’. JCI Insight 3 (6): e97918. 10.1172/jci.insight.97918.

Burgess, Stephen, Christopher N. Foley, and Verena Zuber. 2018. ‘Inferring Causal Relationships Between Risk Factors and Outcomes from Genome-Wide Association Study Data’. Annual Review of Genomics and Human Genetics 19 (August): 303–27. 10.1146/annurev-genom-083117-021731.

Burgess, Stephen, Simon G Thompson, and CRP CHD Genetics Collaboration. 2011. ‘Avoiding Bias from Weak Instruments in Mendelian Randomization Studies’. International Journal of Epidemiology 40 (3): 755–64. 10.1093/ije/dyr036.

Després, Jean-Pierre. 2012. ‘Body Fat Distribution and Risk of Cardiovascular Disease’. Circulation 126 (10): 1301–13. 10.1161/CIRCULATIONAHA.111.067264.

Dewey, Frederick E., Viktoria Gusarova, Colm O’Dushlaine, Omri Gottesman, Jesus Trejos, Charleen Hunt, Cristopher V. Van Hout, et al. 2016. ‘Inactivating Variants in ANGPTL4 and Risk of Coronary Artery Disease’. New England Journal of Medicine 374 (12): 1123–33. 10.1056/NEJMoa1510926.

Ference, Brian A., John J. P. Kastelein, Kausik K. Ray, Henry N. Ginsberg, M. John Chapman, Chris J. Packard, Ulrich Laufs, et al. 2019. ‘Association of Triglyceride-Lowering LPL Variants and LDL-C–Lowering LDLR Variants With Risk of Coronary Heart Disease’. JAMA 321 (4): 364–73. 10.1001/jama.2018.20045.

Ghodsian, Nooshin, Erik Abner, Connor A. Emdin, Émilie Gobeil, Nele Taba, Mary E. Haas, Nicolas Perrot, et al. 2021. ‘Electronic Health Record-Based Genome-Wide Meta-Analysis Provides Insights on the Genetic Architecture of Non-Alcoholic Fatty Liver Disease’. *Cell Reports*. Medicine 2 (11): 100437. 10.1016/j.xcrm.2021.100437.

Gotto, A. M. 1998. ‘Triglyceride as a Risk Factor for Coronary Artery Disease’. The American Journal of Cardiology 82 (9A): 22Q–25Q. 10.1016/s0002-9149(98)00770-x.

Graham, Sarah E., Shoa L. Clarke, Kuan-Han H. Wu, Stavroula Kanoni, Greg J. M. Zajac, Shweta Ramdas, Ida Surakka, et al. 2021. ‘The Power of Genetic Diversity in Genome-Wide Association Studies of Lipids’. Nature 600 (7890): 675–79. 10.1038/s41586-021-04064-3.

Gusarova, Viktoria, Colm O’Dushlaine, Tanya M. Teslovich, Peter N. Benotti, Tooraj Mirshahi, Omri Gottesman, Cristopher V. Van Hout, et al. 2018. ‘Genetic Inactivation of ANGPTL4 Improves Glucose Homeostasis and Is Associated with Reduced Risk of Diabetes’. Nature Communications 9 (1): 2252. 10.1038/s41467-018-04611-z.

Gutgsell, Aspen R., Swapnil V. Ghodge, Albert A. Bowers, and Saskia B. Neher. 2019. ‘Mapping the Sites of the Lipoprotein Lipase (LPL)-Angiopoietin-like Protein 4 (ANGPTL4) Interaction Provides Mechanistic Insight into LPL Inhibition’. The Journal of Biological Chemistry 294 (8): 2678–89. 10.1074/jbc.RA118.005932.

K. Nilsson, Stefan. 2023. ‘First-in-Human Phase I Trial with Lipigons Lipid-Lowering Candidate Lipisense - Lipigon’. 12 July 2023. https://www.lipigon.se/media/pressmeddelanden/first-in-human-phase-i-trial-with-lipigons-lipid-lowering-ca-96244.

Kersten, Sander. 2021. ‘Role and Mechanism of the Action of Angiopoietin-like Protein ANGPTL4 in Plasma Lipid Metabolism’. Journal of Lipid Research 62 (January). 10.1016/j.jlr.2021.100150.

Kurki, Mitja I., Juha Karjalainen, Priit Palta, Timo P. Sipilä, Kati Kristiansson, Kati M. Donner, Mary P. Reeve, et al. 2023. ‘FinnGen Provides Genetic Insights from a Well-Phenotyped Isolated Population’. Nature 613 (7944): 508–18. 10.1038/s41586-022-05473-8.

Landfors, Fredrik, Elin Chorell, and Sander Kersten. 2023. ‘Genetic Mimicry Analysis Reveals the Specific Lipases Targeted by the ANGPTL3-ANGPTL8 Complex and ANGPTL4’. Journal of Lipid Research 64 (1): 100313. 10.1016/j.jlr.2022.100313.

Lichtenstein, Laeticia, Frits Mattijssen, Nicole J. de Wit, Anastasia Georgiadi, Guido J. Hooiveld, Roelof van der Meer, Yin He, et al. 2010. ‘Angptl4 Protects against Severe Proinflammatory Effects of Saturated Fat by Inhibiting Fatty Acid Uptake into Mesenteric Lymph Node Macrophages’. Cell Metabolism 12 (6): 580–92. 10.1016/j.cmet.2010.11.002.

Lipigon Pharmaceuticals, AB. 2023. ‘Lipisense® Progressing to Phase II - What Comes Next?’ 12 December 2023. https://view.news.eu.nasdaq.com/view?id=b012022862ab3ead192d18508d5d82374&lang=en&src=micro.

Liu, Yi, Nicolas Basty, Brandon Whitcher, Jimmy D. Bell, Elena P. Sorokin, Nick van Bruggen, E. Louise Thomas, and Madeleine Cule. 2021. ‘Genetic Architecture of 11 Organ Traits Derived from Abdominal MRI Using Deep Learning’. eLife 10 (June): e65554. 10.7554/eLife.65554.

Mahajan, Anubha, Cassandra N. Spracklen, Weihua Zhang, Maggie C. Y. Ng, Lauren E. Petty, Hidetoshi Kitajima, Grace Z. Yu, et al. 2022. ‘Multi-Ancestry Genetic Study of Type 2 Diabetes Highlights the Power of Diverse Populations for Discovery and Translation’. Nature Genetics 54 (5): 560–72. 10.1038/s41588-022-01058-3.

Manipur, Ichcha, Guillermo Reales, Jae Hoon Sul, Myung Kyun Shin, Simonne Longerich, Adrian Cortes, and Chris Wallace. 2023. ‘CoPheScan: Phenome-Wide Association Studies Accounting for Linkage Disequilibrium’. Preprint. Genetics. 10.1101/2023.06.29.546856.

Mishra, Aniket, Rainer Malik, Tsuyoshi Hachiya, Tuuli Jürgenson, Shinichi Namba, Daniel C. Posner, Frederick K. Kamanu, et al. 2022. ‘Stroke Genetics Informs Drug Discovery and Risk Prediction across Ancestries’. Nature 611 (7934): 115–23. 10.1038/s41586-022-05165-3.

Myocardial Infarction Genetics and CARDIoGRAM Exome Consortia Investigators, Nathan O. Stitziel, Kathleen E. Stirrups, Nicholas G. D. Masca, Jeanette Erdmann, Paola G. Ferrario, Inke R. König, et al. 2016. ‘Coding Variation in ANGPTL4, LPL, and SVEP1 and the Risk of Coronary Disease’. The New England Journal of Medicine 374 (12): 1134–44. 10.1056/NEJMoa1507652.

Pierce, Brandon L, Habibul Ahsan, and Tyler J VanderWeele. 2011. ‘Power and Instrument Strength Requirements for Mendelian Randomization Studies Using Multiple Genetic Variants’. International Journal of Epidemiology 40 (3): 740–52. 10.1093/ije/dyq151.

Richardson, Tom G., Eleanor Sanderson, Tom M. Palmer, Mika Ala-Korpela, Brian A. Ference, George Davey Smith, and Michael V. Holmes. 2020. ‘Evaluating the Relationship between Circulating Lipoprotein Lipids and Apolipoproteins with Risk of Coronary Heart Disease: A Multivariable Mendelian Randomisation Analysis’. PLoS Medicine 17 (3): e1003062. 10.1371/journal.pmed.1003062.

Singh, Abhishek K., Balkrishna Chaube, Xinbo Zhang, Jonathan Sun, Kathryn M. Citrin, Alberto Canfrán-Duque, Binod Aryal, et al. 2021. ‘Hepatocyte-Specific Suppression of ANGPTL4 Improves Obesity-Associated Diabetes and Mitigates Atherosclerosis in Mice’. Journal of Clinical Investigation 131 (17): e140989. 10.1172/JCI140989.

Sinnott-Armstrong, Nasa, Isabel S. Sousa, Samantha Laber, Elizabeth Rendina-Ruedy, Simon E. Nitter Dankel, Teresa Ferreira, Gunnar Mellgren, et al. 2021. ‘A Regulatory Variant at 3q21.1 Confers an Increased Pleiotropic Risk for Hyperglycemia and Altered Bone Mineral Density’. Cell Metabolism 33 (3): 615–628.e13. 10.1016/j.cmet.2021.01.001.

Stanzick, Kira J., Yong Li, Pascal Schlosser, Mathias Gorski, Matthias Wuttke, Laurent F. Thomas, Humaira Rasheed, et al. 2021. ‘Discovery and Prioritization of Variants and Genes for Kidney Function in >1.2 Million Individuals’. Nature Communications 12 (1): 4350. 10.1038/s41467-021-24491-0.

Sveinbjornsson, Gardar, Magnus O. Ulfarsson, Rosa B. Thorolfsdottir, Benedikt A. Jonsson, Eythor Einarsson, Gylfi Gunnlaugsson, Solvi Rognvaldsson, et al. 2022. ‘Multiomics Study of Nonalcoholic Fatty Liver Disease’. Nature Genetics 54 (11): 1652–63. 10.1038/s41588-022-01199-5.

The Gtex Consortium. 2020. ‘The GTEx Consortium Atlas of Genetic Regulatory Effects across Human Tissues’. Science 369 (6509): 1318–30. 10.1126/science.aaz1776.

Wallace, Chris. 2021. ‘A More Accurate Method for Colocalisation Analysis Allowing for Multiple Causal Variants’. PLoS Genetics 17 (9): e1009440. 10.1371/journal.pgen.1009440.

Ward, Lucas D., Ho-Chou Tu, Chelsea B. Quenneville, Shira Tsour, Alexander O. Flynn-Carroll, Margaret M. Parker, Aimee M. Deaton, et al. 2021. ‘GWAS of Serum ALT and AST Reveals an Association of SLC30A10 Thr95Ile with Hypermanganesemia Symptoms’. Nature Communications 12 (1): 4571. 10.1038/s41467-021-24563-1.

Yin, Wu, Stefano Romeo, Shurong Chang, Nick V. Grishin, Helen H. Hobbs, and Jonathan C. Cohen. 2009. ‘Genetic Variation in ANGPTL4 Provides Insights into Protein Processing and Function*’. Journal of Biological Chemistry 284 (19): 13213–22. 10.1074/jbc.M900553200.

Zhou, Wei, Jonas B. Nielsen, Lars G. Fritsche, Rounak Dey, Maiken E. Gabrielsen, Brooke N. Wolford, Jonathon LeFaive, et al. 2018. ‘Efficiently Controlling for Case-Control Imbalance and Sample Relatedness in Large-Scale Genetic Association Studies’. Nature Genetics 50 (9): 1335–41. 10.1038/s41588-018-0184-y.

Zou, Yuxin, Peter Carbonetto, Gao Wang, and Matthew Stephens. 2022. ‘Fine-Mapping from Summary Data with the “Sum of Single Effects” Model’. PLOS Genetics 18 (7): e1010299. 10.1371/journal.pgen.1010299.

Zuber, Verena, Dipender Gill, Mika Ala-Korpela, Claudia Langenberg, Adam Butterworth, Leonardo Bottolo, and Stephen Burgess. 2021. ‘High-Throughput Multivariable Mendelian Randomization Analysis Prioritizes Apolipoprotein B as Key Lipid Risk Factor for Coronary Artery Disease’. International Journal of Epidemiology 50 (3): 893– 901. 10.1093/ije/dyaa216.

